# The Hybrid Forecasting Method SVR-ESAR for Covid-19

**DOI:** 10.1101/2020.05.20.20103200

**Authors:** Juan Frausto Solis, José Enrique Olvera Vazquez, Juan J González Barbosa, Guadalupe Castilla Valdez, Juan Paulo Sánchez Hernández, Joaquín Perez-Ortega

**Affiliations:** Tecnológico Nacional de Mexico, IT Cd Madero; Universidad Politécnica del Estado de Morelos; Tecnológico Nacional de Mexico, CENIDET

## Abstract

We know that SARS-Cov2 produces the new COVID-19 disease, which is one of the most dangerous pandemics of modern times. This pandemic has critical health and economic consequences, and even the health services of the large, powerful nations may be saturated. Thus, forecasting the number of infected persons in any country is essential for controlling the situation. In the literature, different forecasting methods have been published, attempting to solve the problem. However, a simple and accurate forecasting method is required for its implementation in any part of the world. This paper presents a precise and straightforward forecasting method named SVR-ESAR (Support Vector regression hybridized with the classical Exponential smoothing and ARIMA). We applied this method to the infected time series in four scenarios, which we have taken for the Github repository: the Whole World, China, the US, and Mexico. We compared our results with those of the literature showing the proposed method has the best accuracy.

## 1. Introduction

In December 2019, Chinese citizens started to suffer a strange respiratory disease; this happened in Wuhan city, Hubei province. This disease was produced by a virus named SARS-CoV-2 or COVID-19, whose origin remains unknown [1], [2]. However, the scientists observed this disease is very contagious. It spread rapidly throughout the world, becoming a pandemic with an increased death rate as it left China. Thus, knowing the behavior of the disease is of utmost importance to take the appropriate sanitary measures. Researchers observed the pandemic rules in different countries; they noticed that when the control measures were relaxed, and a large number of people severely infected, the health services could be saturated. Therefore, it is necessary to establish efficient methods for estimating the number of infected people accurately; thus, this helps the health authorities to take the correct measures at the appropriate time using SIR or other pandemic models. There are two main approaches for describing an epidemic:

- The SIRs models. The mathematical epidemic models, the first of which was proposed by Sir Roland Ross in 1902 and adjusted in 1927 by Kermack and McKendrick [3].
- Forecasting models using time series. This approach is an ancient method for describing an epidemic such as Covid-19. For instance, [4] shows different forecasting epidemic studies using time series.

These two main epidemic approaches are fundamental, and they are related. However, the infected prediction of time series is one of the biggest challenges since the estimations require the lowest error. Currently, the forecasting time series techniques can be classified as:

- Classical forecasting ARIMA and Exponential Smoothing (ES) and they typically obtain good results [4],
- Artificial Intelligence techniques where vector support machines (SVM) and neural networks are the most common.

Nowadays, it is common to use hybrid methods using these two techniques. For instance, [5] combines ES and ARIMA. Roughly speaking, it is a two-phases forecasting technique. Once the first phase obtains a first approximation forecast (or base forecast), a second phase improves the latter by using the residual values of the first step. Koning et al. and Makridakis et al. presented this approach for time series from M3-competition with very good results [6], [7]. Also, among the best methods for M4-competition are hybrid methods such as SVR with ES and ARIMA [8]. Inspired by these ideas, we decided to design a forecasting hybrid method for the Covid-19 time series. This method is named SVR-ESAR (SVR with ES and ARIMA), and it is presented in section three, after a brief background in section two. Then we used infected cases that we have taken from the Github repository (https://github.com/CSSEGISandData/COVID19/blob/master/archived_data/archived_time_series/time_series_19-covid-Confirmed_archived_0325.csv) and we prepared foiur scenarios for and experimentation is described in section four. Finally, we present the conclusions in section five.

## 2. Background

Epidemics have always existed in the world. However, their first model (or SIR model) was not published until 1902 [3]. This model and its variants are based on Markov models, where the population is divided into several classes as Susceptible, Infected (or confirmed), and Recovered. This paper is focused on forecasting the confirmed class by using time series. We use a hybrid method that uses Support Vector Regression (SVR), Exponential Smoothing ES), and ARIMA. These techniques are widespread in several platforms, and they are briefly explained as follows:

- Support Vector Regression (SVR): This is a very effective method for forecasting time series, which is an application of Vector Support Machine (SVM) proposed by Vapnik [9]. SVR and SVM minimize the margin error and use Kernel function for non-separable classes. For obtaining good forecasting results, in the quality and stability of SVR, its parameters and the kernel parameters should be tuned. Typically, they are tuned by a grid search, a genetic or other heuristic selects the best parameters [10].
- **Exponential smoothing (ES)**. This forecasting method uses an exponentially weighted moving average with the values of the times series previously observed. ES smooths (averages) the past values of the time series with the last forecast. Exponential smoothing has given good results in different forecasting competitions, especially for short series [11], [12].
- ARIMA model: This model is described in terms of parameters currently named p, d, and q, and linked to three types of processes: autoregression, integration, and the moving average [13].

## 3. Description of the SVR-ESAR method

We developed a hybrid Forecasting method named SVR-ESAR, which uses GA-SVR and an adjustment phase. The SVR-ESAR Architecture GA-SVR has two phases and is shown in Figure 1:

**Figure 1.**
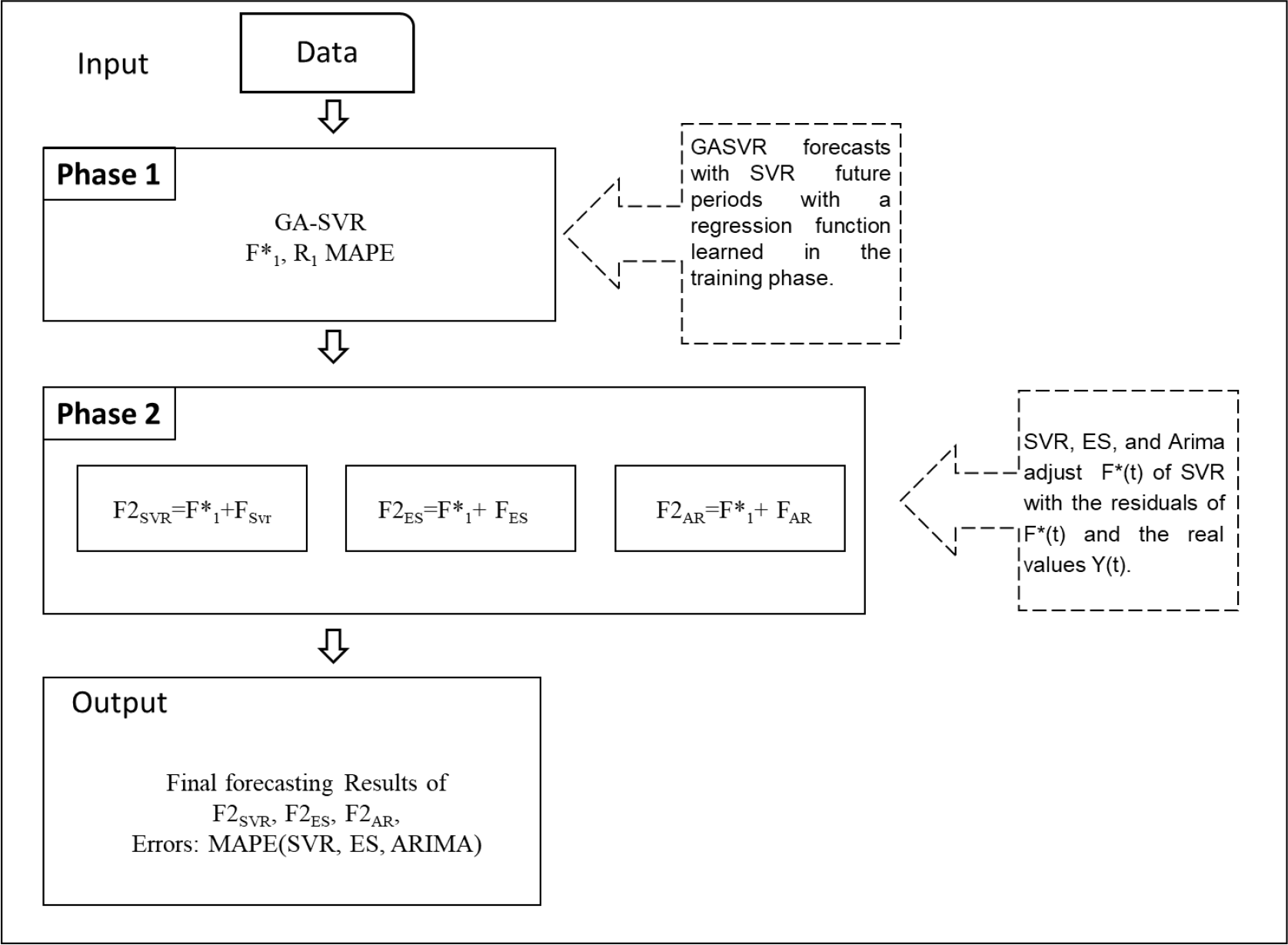
SVR-ESAR Architecture.

❖ Phase 1: A Genetic Algorithm adjusts the parameters of an SVR machine and its kernels (linear, rbf, sigmoid) using Grid Search and iteratively improves the F(t) forecast of SVR [10]. The improved forecast F*_1_(t) in this phase has a MAPE error dependent on the residuals during the training phase.
❖ Phase 2: Three alternative techniques obtain a correction of the F*_1_(t) forecast previously obtained by GA-SVR in phase 1: SVR, Holt Exponential Smoothing (ES), and ARIMA [13]. These techniques are applied to the residuals between F*_1_(t) and the actual values in the training stage, and obtaining a better forecasting F2svr, F2_ES_, and F2_AR_ with the application in the second phase of these three techniques SVR, ES, and ARIMA respectively.

## 4. Experimental Results

The presented forecasting method was applied to people infected with Coronavid 19. The period for this time series is from 22/January until 25/April/ 2020 [14]; we have taken the data from the GitHub repository. In the references [4] and [13], the forecast is for the last ten days. Thus, we used the same parameters to compare their results with our proposed forecasting method. We have four scenarios:

- The Whole World: The forecasting results obtained for infected people [15]. We compare the results obtained for our proposed forecasting method using the infected cases.
- China: This country was the origin of the pandemic. There are published results for this case [4], which we use in this section for validating the proposed method.
- United States (US): This country has the highest number of incidences in the American continent; also, it is significant because it has a lot of communication with the rest of the world.
- Mexico implemented some particular policies and has a lot of relationships with many countries, particularly with the US, and Canada.

Table 1 presents the results for the Whole World and China, obtained with the application of the proposed method for confirmed forecasting case. In this table, we show the results of SVR-ESAR using the adjustment techniques SVR, ES, and ARIMA. We notice that all these techniques have a small value of MAPE error. We also present the results obtained for this case in [4] and [15]. We observe that SVR-ARIMA achieved the best results for these cases. Furthermore, Figure 2 shows the results obtained by SVR-ESAR for these scenarios.

**Table 1.** Forecasting for the infected in the Whole World and China using the SVR-ESAR method with three adjustment techniques (SVR, Exponential Smoothing, and ARIMA)

| Scenario 1: Infected in the Whole Word. |  |  |  |  |
| --- | --- | --- | --- | --- |
| Forecasting method | Exponential Smoothing, [15] | SVR-ESAR |  |  |
|  | Adjusted Technique: | Adjusted Technique |  |  |
|  | Multiplicative Trend model | SVR | ES | Arima |
| MAPE error (%) | 6.9 | 2.46 | 0.77 | 0.56 |
| Scenario 2: Infected in China |  |  |  |  |
| Forecasting method | Neural Networks [4] | SVR-ESAR |  |  |
|  | Adjusted Technique | Adjusted Technique |  |  |
|  | Unknown | SVR | ES | Arima |
| MAPE error (%) | 4.79 | 1.90 | 3.53 | 1.41 |

**Figure 2:**
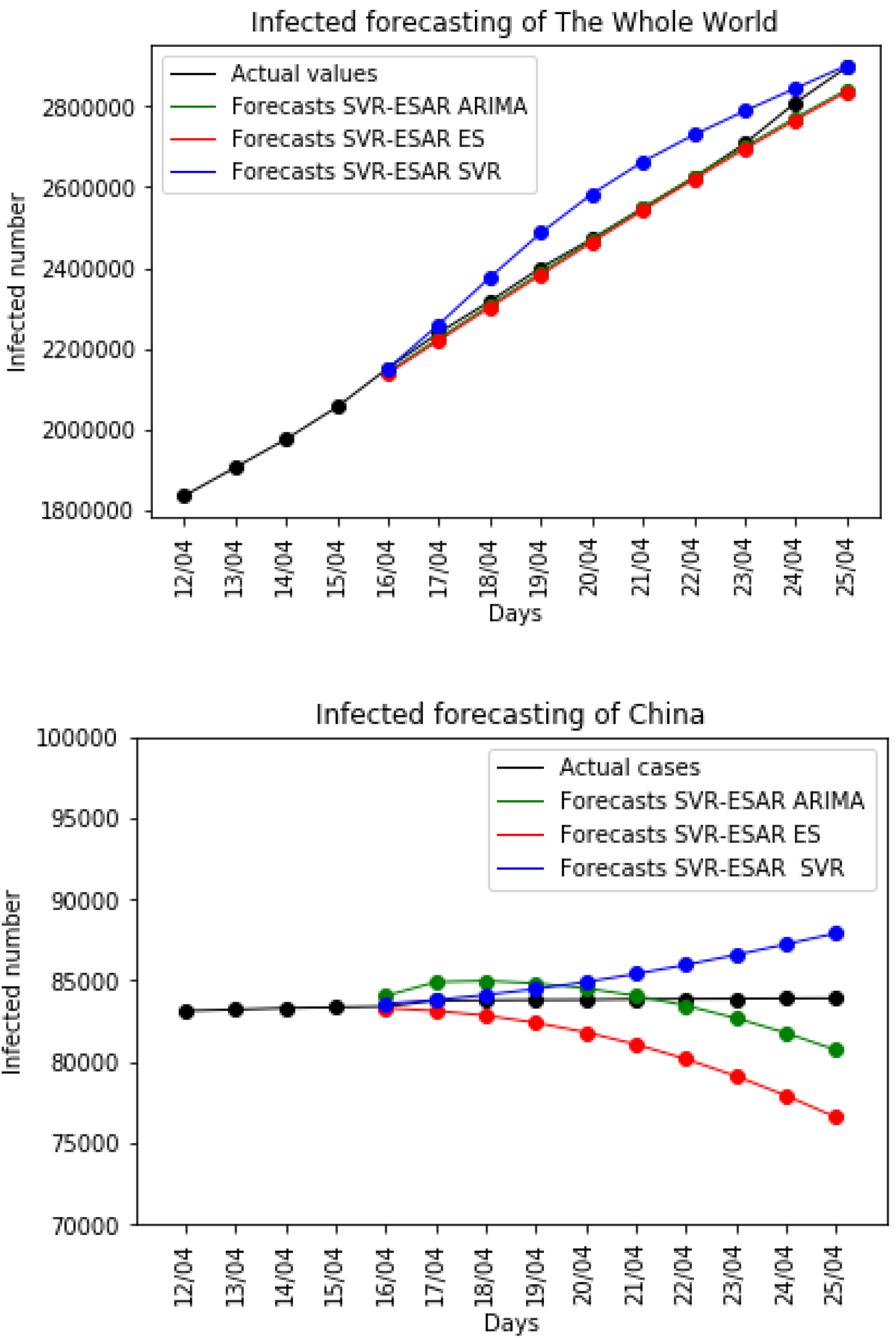
Forecast of SVR-ESAR for the Whole World and China

We present in Table 2 the forecasting confirmed results for the US and Mexico obtained with SVR-ESAR. We show the results of the proposed method using the adjustment obtained by SVR, ES, and ARIMA (Figure 3). We point out that in the case of the US, the ARIMA adjustment technique achieves the best result. This table also shows the results obtained by SVR-ESAR for Mexico. This time the proposed method achieves a modest forecasting result with all three adjustment techniques.

**Table 2.** Forecasting for the infected in the Whole World and China using the SVR-ESAR method with three adjustment techniques (SVR, Exponential Smoothing, and ARIMA)

| Scenario 3 and 4: Infected in United States and Mexico |  |  |  |
| --- | --- | --- | --- |
| Forecasting Method | SVR-ESAR |  |  |
|  | Adjusted Technique |  |  |
|  | SVR | ES | Arima |
| MAPE US (%) | 5.09 | 5.39 | 0.90 |
| MAPE Mexico (%) | 12.44 | 13.25 | 12.57 |

**Figure 3.**
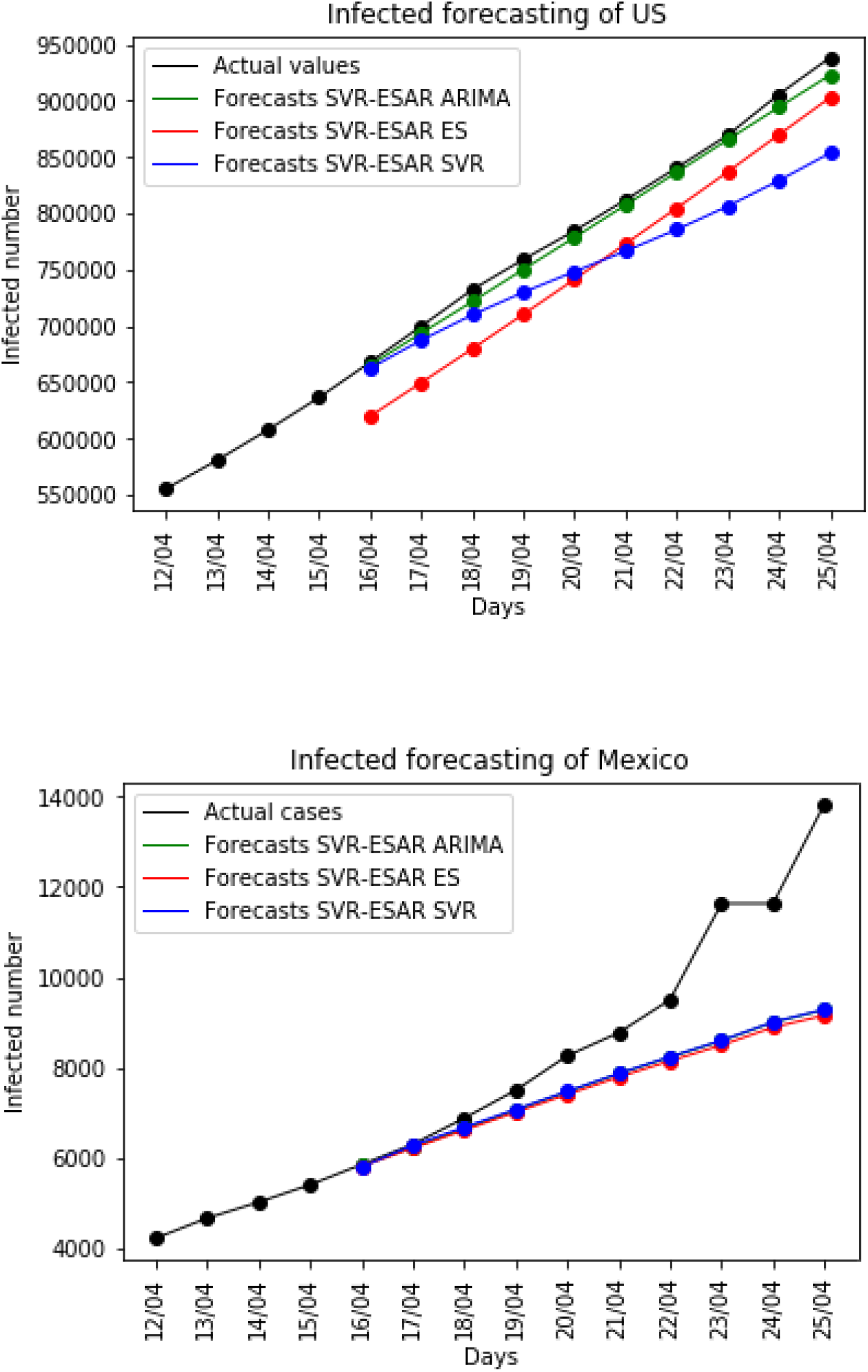
The forecasting of infected cases for the US and Mexico

## 5. Discussion and conclusion

This paper presents a forecasting method named SVR-ESAR (Support Vector Regression with Exponential Smoothing and ARIMA), which was applied for the estimation of confirmed (infected) cases of Covid-19 in four scenarios (the Whole World, China, the US, and Mexico). This method is straightforward and uses SVR with an iterative adjustment phase. In the adjustment phase, we applied three algorithms to the SVR residuals: ES, SVR, and ARIMA. We found SVR-ESAR with an ARIMA adjustment obtained the best or equivalent results for all these scenarios. We applied this method for data of Mexico with modest quality results. However, the results of SVR-ESAR were compared with those published in the literature for the US and China, and we found the proposed method achieved the best predictions.

## Data Availability

All the data used in this study are described in the manuscript. We have not used clinical trials and neither any other prospective interventional study.

https://github.com/CSSEGISandData/COVID19/blob/master/archived_data/archived_time_series/time_series_19-covid-Confirmed_archived_0325.csv

## Authors contribution

**Conceptualization:** Juan Frausto Solís.

**Data curation:** José Enrique Olvera Vázquez and Javier Gonzalez Barbosa.

**Investigation:** Juan Frausto Solis.

**Methodology:** Guadalupe Castillo Valdez.

**Project Supervisor:** Juan Frausto Solís.

**Software:** Juan Paulo Sánchez, Guadalupe Castillo, José Enrique Olvera Vázquez.

**Validation:** Javier González Barbosa.

**Writing – original draft:** Juan Frausto Solís, Javier Gonzalez Barbosa.

**Writing – review & editing:** Joquin Perez Ortega.

